# Pre-pandemic mental health and disruptions to healthcare, economic, and housing outcomes during COVID –19: evidence from 12 UK longitudinal studies

**DOI:** 10.1101/2021.04.01.21254765

**Authors:** Giorgio Di Gessa, Jane Maddock, Michael J. Green, Ellen J. Thompson, Eoin McElroy, Helena L. Davies, Jessica Mundy, Anna J. Stevenson, Alex S.F. Kwong, Gareth J. Griffith, Srinivasa Vittal Katikireddi, Claire L. Niedzwiedz, George B. Ploubidis, Emla Fitzsimons, Morag Henderson, Richard J. Silverwood, Nish Chaturvedi, Gerome Breen, Claire J. Steves, Andrew Steptoe, David J Porteous, Praveetha Patalay

## Abstract

**Background:** The COVID-19 pandemic and associated virus suppression measures have disrupted lives and livelihoods and people already experiencing mental ill-health may have been especially vulnerable.

**Aim:** To quantify mental health inequalities in disruptions to healthcare, economic activity and housing.

**Method:** 59,482 participants in 12 UK longitudinal adult population studies with data collected prior to and during the COVID-19 pandemic. Within each study we estimated the association between psychological distress assessed pre-pandemic and disruptions since the start of the pandemic to three domains: healthcare (medication access, procedures, or appointments); economic activity (employment, income, or working hours); and housing (change of address or household composition). Meta-analyses were used to pool estimates across studies.

**Results:** Across the analysed datasets, one to two-thirds of participants experienced at least one disruption, with 2.3-33.2% experiencing disruptions in two or more domains. One standard deviation higher pre-pandemic psychological distress was associated with: (i) increased odds of any healthcare disruptions (OR=1.30; [95% CI:1.20–1.40]) with fully adjusted ORs ranging from 1.24 [1.09–1.41] for disruption to procedures and 1.33 [1.20– 1.49] for disruptions to prescriptions or medication access; (ii) loss of employment (OR=1.13 [1.06–1.21]) and income (OR=1.12 [1.06 –1.19]) and reductions in working hours/furlough (OR=1.05 [1.00–1.09]); (iii) no associations with housing disruptions (OR=1.00 [0.97–1.03]); and (iv) increased likelihood of experiencing a disruption in at least two domains (OR=1.25 [1.18–1.32]) or in one domain (OR=1.11 [1.07–1.16]) relative to no disruption.

**Conclusion:** People experiencing psychological distress pre-pandemic have been more likely to experience healthcare and economic disruptions, and clusters of disruptions across multiple domains during the pandemic. Failing to address these disruptions risks further widening the existing inequalities in mental health.

## Introduction

The COVID-19 pandemic and consequent mitigation measures have led to notable changes to routine healthcare delivery, economic participation, and housing circumstances in many countries. There is extensive evidence that the negative impacts of the pandemic disproportionately affect certain socio-demographic groups (e.g., socio-economically disadvantaged, ethnic minorities, younger generations, and women) (1). However, although poor mental health might be an important indicator of inequity in these outcomes, to date little is known about whether individuals with poor mental health are at particular risk of these disruptions during the pandemic (2, 3).

Mental health conditions like depression and anxiety are widespread in the population with one in six adults estimated to suffer from these conditions at any given time (4). People with prior mental health difficulties have experienced higher risk for COVID-19 related adverse outcomes including greater risk of infection, severe disease, and mortality (5). In addition, these individuals had already experienced greater risk of social and health inequalities prior to the pandemic (6, 7). Moreover, recent evidence suggests they are less likely to be vaccinated, further increasing the risk of infection-related adverse outcomes for this group (8). There has been less attention paid to whether non-infection related outcomes of the pandemic - such as healthcare, economic, and housing disruptions – have been differentially experienced by those with poor mental health. Evidence from previous disruptive events, such as economic recessions, highlights greater negative consequences for those with poor mental health (9).

This study investigates the extent to which pre-pandemic psychological distress (symptoms of anxiety and depression) was associated with experiences of healthcare, economic, and housing disruptions in the UK during the COVID-19 pandemic. We examine whether this association differs between socio-demographic groups based on sex, age, ethnicity, and socio-economic position. We also examine the prevalence of, and associations with disruptions across multiple domains, as people who face adverse disruptions in multiple domains are likely to have poorer longer-term outcomes. We use data from over 59,000 participants across 12 UK population-based longitudinal studies with rich pre-pandemic socio-demographic and health measures as well as detailed information about disruptions during the pandemic.

## Methods

### Design

The UK National Core Studies – Longitudinal Health and Wellbeing initiative aims to co-ordinate primary analyses across multiple UK longitudinal population-based studies (https://www.ucl.ac.uk/drupal/site_covid-19-longitudinal-health-wellbeing/). Even with the same research question and data source, research has highlighted that results can vary due to methodological heterogeneity and researcher decisions (10). In this programme of work, by conducting analyses in a co-ordinated manner across different datasets, we minimise such biases and maximise comparability, while appropriately accounting for the study design and characteristics of individual datasets. Synthesis of findings across studies allows pooling of evidence across a larger sample size, including subgroup analyses by age and other socio-demographic groups (e.g., sex, ethnicity).

### Participants

Data were drawn from 12 UK population studies which conducted surveys both before and during the COVID-19 pandemic. Details of the design, sample frames, current age range, timing of the most recent pre-pandemic and COVID-19 surveys, response rates, and analytical sample size are available in Table 1. Demographic and socio-economic characteristics of each analytical sample are presented in Table S1.

**Table 1.** Details of each included study.

| Study Population | Design and Sample Frame | 2020 Age Range in years | Most recent pre-pandemic survey | Details of 2020 covid surveys (response rate) | Analytic N |
| --- | --- | --- | --- | --- | --- |
| *Age Homogenous Cohorts* |  |  |  |  |  |
| MCS: Millennium Cohort Study | Cohort of UK children born between Sept 2000 and Jan 2002 with regular follow-up surveys from birth. | 18-20 | 2018 | Two surveys: May (26.6%) & Sep-Oct (24.2%) | 3028 |
| ALSPAC (G1): Avon Longitudinal Study of Parents and Children-Generation 1 | Cohort of children born in the South-West of England between April 1991 and Dec 1992, with regular follow-up questionnaires from birth (original young people). | 27-29 | 2017-2018 | Three questionnaires: April (19%), June (17.4%), December (26.4%) | 2698 |
| NS: Next Steps, formerly known as Longitudinal Study of Young People in England | Sample recruited via secondary schools in England at around age 13 with regular follow-up surveys thereafter. | 29-31 | 2015 | Two surveys: May (20.3%) & Sep-Oct (31.8%) | 3209 |
| BCS70: British Cohort Study 1970 | Cohort of all children born in Great Britain (i.e. England, Wales & Scotland) in one week in 1970, with regular follow-up surveys from birth. | 50 | 2016 | Two surveys: May (40.4%) & Sep-Oct (43.9%) | 4303 |
| NCDS: National Child Development Study | Cohort of all children born in Great Britain (i.e. England, Wales & Scotland) in one week in 1958, with regular follow-up surveys from birth. | 62 | 2013 | Two surveys: May (57.9%) & Sep-Oct (53.9%) | 5394 |
| NSHD: National Survey of Health and Development | Cohort of all children born in Great Britain (i.e. England, Wales & Scotland) in one week in 1946, with regular follow-up surveys from birth. | 74 | 2015 | Two surveys: May (68.2%) & Sep-Oct (61.5%) | 1310 |
| *Age Heterogeneous Studies* |  |  |  |  |  |
| USOC: Understanding Society: the UK Household Longitudinal Survey | A nationally representative longitudinal household panel study, based on a clustered-stratified probability sample of UK households, with all adults aged 16+ in chosen households surveyed annually. | 16-96 | 2018-2019 | Six: surveys: April (40.3%), May (33.6%), Jun (32.0%), July (31.2%), Sep (29.2%) & Nov (27.3%) | 13175 |
| ELSA: English Longitudinal Study of Aging | A nationally-representative population study of individuals aged 50+ living in England, with biennial surveys and periodic refreshing of the sample to maintain representativeness. | 52-90+ | 2018-2019 | Two surveys: Jun-July (75%) & Nov-Dec (73%) | 5061 |
| GS: Generation Scotland: the Scottish Family Health Study | A family-structured, population-based Scottish cohort, with participants aged 18-99 recruited between 2006-2011 | 27-100 | 2006-2011 | Two surveys: April-Jun (21.6%) & Jul-Aug (15.6%) | 3179 |
| ALSPAC(G0): Avon Longitudinal Study of Parents and Children-Generation 0 | Parents of the ALSPAC(G1) cohort described above, treated as a separate age-heterogenous study population. (original parents) | 45-81 | 2011-2013 | Three questionnaires: April (12.4%), June (12.2%), December (14.3%) | 3212 |
| TWINSUK: the UK Adult Twin Registry | A cohort of volunteer adult twins (55% monozygotic and 43% dizygotic) from around the United Kingdom who were sampled between 18-101 years of age. | 22-96 | 2017-2018 | Three surveys: April (64.3%), July (77.6%) & November | 2855 |
|  |  |  |  | (76.1%) |  |
| Genetic Links to Anxiety and Depression (GLAD) study | Participants with depression and/or anxiety aged 16+ from the 2018 Genetic Links to Anxiety and Depression study (GLAD) were invited to take part in covid surveys as part of a new project, Covid-19 Psychiatry and Neurological Genetics study (COPING). | 16-89 | 2018-2021 (data from GLAD) | Fortnightly data collection from April to July (20.4%), then monthly (19,7%). | 12107 |

Six of these were age homogenous birth cohorts (all individuals similar age): the Millennium Cohort Study (MCS; 11); the Avon Longitudinal Study of Parents and Children (ALSPAC G1; 12); Next Steps (NS, formerly known as the Longitudinal Study of Young People in England; 13); the 1970 British Cohort Study (BCS; 14), the National Child Development Study (NCDS; 15); and the National Survey of Health and Development (NSHD; 16).

Six studies covered a range of ages. These age heterogenous studies were: Understanding Society (USoc; 17); the English Longitudinal Study of Ageing (ELSA; 18); Generation Scotland: the Scottish Family Health Study (GS; 19, 20); the UK Adult Twin Registry (TWINS; 21); the Genetic Links to Anxiety and Depression study (GLAD; 22), which is a cohort of those with experience of anxiety and/or depression; and the parents of the ALSPAC-G1 cohort (ALSPAC-G0)(23).

Analytical samples included those who had a measure of psychological distress in a recent pre-pandemic survey, had information available for at least one outcome in a COVID-19 survey, and had valid data on a minimum set of covariates (sex, ethnicity, socio-economic position, and age). Each study was weighted to be representative of its target population, accounting for sampling design, attrition up to the most recent pre-pandemic survey, and differential non-response to the COVID-19 surveys.

Ethics approvals were received for data collection in all studies and the specifics for each included study are detailed in supplementary file 2.

### Measures

Below we describe the overall approach to measuring each variable in the analysis. Full details of the questions and coding used within each cohort are available in supplementary file 1.

### Exposure: Pre-pandemic psychological distress

All studies measured psychological distress in the most recent pre-pandemic survey using validated continuous scales. These included GHQ-12 in NS and USOC; GHQ-28 for NSHD and GS; Malaise Inventory in NCDS and BCS70; K-6 in MCS; and CES-D in ELSA. Table S2 presents details of the measure in each study, including when last collected, its distribution (mean, range, and standard deviation), and the percentage with high psychological distress in each study. Pre-pandemic measures of distress had been taken some years prior to the pandemic (median 3.3 years; interquartile range: 1.9-6.5 years). Each scale was transformed into standard deviation units (z-scores) within each cohort, and we conducted additional analyses using dichotomous variables based on established cut-offs for each measure.

### Outcomes

Outcomes were disruptions separated into three broad domains: healthcare, economic, and housing. For healthcare, we assessed any reported disruptions to: prescriptions or medication access; procedures or surgery; and appointments (e.g., with a GP or outpatient services). Any deviation from planned/existing treatment was coded as a disruption, regardless of the reason for the disruption. In the economic domain we assessed disruptions to: usual economic activity (i.e., education/training, occupations); job loss; loss of income; or any changes in working hours, including furlough. Housing disruptions included: any loss of housing or change of address; and any changes in household composition (i.e., who the participant lives with). We generated variables indicating any disruption within each domain, and the number of domains in which disruptions had occurred: no disruptions, disruption in one domain, or disruptions in two or more domains. Where multiple survey waves had been conducted during the pandemic, we produced a single variable indicating any relevant disruption reported up to and including the most recent survey. Most studies had at least seven months of follow-up after the start of the pandemic in March 2020 (see Table 1 for details).

### Other variables

All covariates were based on pre-pandemic assessments. We explored subgroup differences by sex (female, male), ethnicity (White, non-White ethnic minority; in cohorts where possible), socio-economic position measured by highest education level (degree, no-degree) and age (16-24; 25-34; 35-44; 45-54; 55-64; 65-74; 75+). Age homogeneous cohorts were included in their corresponding age band.

The following covariates were included where relevant and available within each study: UK nation (i.e., England, Scotland, Wales, or Northern Ireland); partnership status (single or couple); presence of children in the household; housing tenure (owned/mortgage or rented/other); own occupational class (or parental occupational class for younger cohorts; four categories: managerial/professional; intermediate; routine; or never worked/not available/long-term non-employed); prior chronic conditions or illness (yes or no); and an indicator of physical disability (yes or no).

### Analysis

Within each study, the association between each binary disruption outcome and standardised pre-pandemic psychological distress was examined using logistic regression models. In order to examine whether poor mental health is associated with disruptions above and beyond well-known socio-demographic and health characteristics, in multivariable analyses we controlled for a range of factors. Following unadjusted associations, first we adjusted for a common set of covariates across all studies, including, where relevant: age, sex, ethnicity, education, and UK nation (adjustment 1). Second, we further accounted for relevant prior health and other relevant confounders such as partnership status, presence of children, housing tenure, occupational class, prior chronic conditions, and physical disability (adjustment 2). For this additional adjustment, variables were created to be as comparable as possible across studies while being suitable for cohort-specific characteristics. Sub-group differences were explored with stratified regressions predicting any disruption in each domain, and the minimal adjustment set (for optimal comparability across studies). Details of all these measures and how they were assessed in each study are presented in supplementary file 1. As an additional sensitivity analysis, the non-stratified models predicting any disruption in each domain were repeated using established categorical cut-offs (reflecting high psychological distress symptoms) as the exposure. Details of the measure-specific cut-off points used are available in Supplementary File 2.

Results from each study were then pooled for each outcome across the studies overall and then stratified by sex, education level, ethnicity, and age. We used a random effects meta-analysis with restricted maximum likelihood. We report heterogeneity using the I^2^ statistic (24). We used random effects meta-regression to investigate whether the between-study heterogeneity could be explained by the time since pre-pandemic mental health measure categorised as: ≤2 years; 2-5 years; 5-7 years; and 7+ years. Meta-analyses were conducted in Stata 16 (25).

## Results

### Descriptive Statistics

Between 7% (TWINS) and 24% (NS) of participants from the population-based cohorts and 54% of participants in GLAD (reflecting their recruitment of those with mental health difficulties) reported high psychological distress prior to the pandemic. As expected, the prevalence of psychological distress was generally higher among women, those without a degree, and younger age groups (see supplementary Tables S3-S4 for full percentages of individuals classified as having high psychological distress, stratified by socio-demographic characteristics). Table 2 shows the percentage of respondents who reported disruptions: this ranged from <10% (MCS, GLAD and TWINSUK) to 37% (ELSA) for healthcare; from 10% (NSHD) to 51% (USOC) for the economic domain; and from 2% (NSHD) to 36% (MCS) for housing. Between 28% (NSHD) and 77% (USOC) of study participants experienced at least one of these disruptions during the pandemic (see supplementary Table S5 for the percent prevalence of any healthcare, economic, and housing disruptions during the pandemic by sex, ethnicity, education level, and age-group).

**Table 2.** Percent prevalence (and 95% confidence intervals) of any healthcare, economic, and housing disruptions during the pandemic as well as of cumulative disruptions, by study.

|  | **MCS** | **ALSPAC G1** | **NS** | **BCS70** | **NCDS** | **NSHD** | **USOC** | **ELSA** | **GS** | **ALSPAC G0** | **TWINS UK** | **GLAD** |
| --- | --- | --- | --- | --- | --- | --- | --- | --- | --- | --- | --- | --- |
| *Based on data until* | *Oct 2020* | *Jan 2021* | *Oct 2020* | *Oct 2020* | *Oct 2020* | *Oct 2020* | *Nov 2020* | *Dec 2020* | *Sept 2020* | *Jan 2021* | *Nov 2020* | *Jan 2021* |
| **Any healthcare disruption** | 9.6<br>(7.9-11.5) | 15.9<br>(14.3-17.6) | 11.6<br>(9.4-14.3) | 13.3<br>(11.7-15.1) | 15.3<br>(13.9-16.8) | 18.5<br>(14.7-22.9) | 31.9<br>(30.8-32.9) | 36.7<br>(34.9-38.4) | 27.4<br>(25.9-29.0) | 19.9<br>(18.1-21.9) | 8.7*<br>(7.7 - 9.8) | 0.7<br>(0.6 - 0.9) |
| Prescription/medication access | 3.3<br>(2.6-4.2) | NA | 3.8<br>(2.5-5.7) | 3.3<br>(2.6-4.3) | 2.5<br>(1.9-3.2) | 2.3<br>(1.3-4.2) | 5.6<br>(5.1-6.2) | 0.8<br>(0.5-1.3) | 6.7<br>(5.8-7.6) | NA | 2.9<br>(2.5- 3.4) | 0.7<br>(0.6 - 0.9) |
| Procedures or surgery | 1.0<br>(0.5-2.2) | 1.6<br>(1.2-2.1) | 1.3<br>(0.5-3.5) | 0.8<br>(0.5-1.0) | 2.1<br>(1.6 - 2.8) | 2.4<br>(1.3-4.5) | 12.3<br>(11.6-13.1) | 21.4<br>(20.0-22.9) | 2.9<br>(2.4-3.6) | 2.9<br>(2.1-3.9) | NA | NA |
| Appointments | 6.4<br>(5.1-8.1) | 11.7<br>(10.3-13.2) | 7.2<br>(5.6-9.1) | 10.2<br>(8.8-11.8) | 12.0<br>(10.7-13.2) | 14.3<br>(10.9-18.4) | 28.5<br>(27.5-29.5) | 21.3<br>(19.8-22.9) | 22.0<br>(20.6-23.5) | 14.4<br>(12.8-16.2) | NA | NA |
| **Any economic disruption** | 43.6<br>(40.5-46.8) | 50.2<br>(47.6-52.9) | 41.0<br>(37.8-44.3) | 40.8<br>(38.4-43.2) | 36.8<br>(35.1-38.6) | 10.0<br>(6.7-14.8) | 51.5<br>(50.3-52.7) | 30.2<br>(28.5-32.0) | 20.8<br>(19.4-22.2) | 48.6<br>(46.2-51.1) | 30.9<br>(29.2-32.6) | 41.9<br>(41.0 - 42.8) |
| Main economic activity | 35.1<br>(31.2-39.2) | 43.3<br>(40.6-46.0) | 7.9<br>(5.7-10.7) | 5.9<br>(4.9-7.0) | 6.6<br>(5.8-7.4) | 0.9<br>(0.5-1.5) | n/a | 4.6<br>(3.8-5.6) | 10.3<br>(9.3-11.4) | 46.6<br>(43.7-49.4) | 17.6<br>(16.6 - 18.6) | 41.0<br>(40.1 - 41.9) |
| Employment | 8.3<br>(6.0-11.3) | 6.4<br>(5.1-7.9) | 7.4<br>(5.3-10.3) | 5.8<br>(4.9-6.9) | 6.3<br>(5.6-7.2) | 0.7<br>(0.4-1.3) | 6.1<br>(5.6-6.7) | 1.4<br>(1.0-2.0) | 0.2<br>(0.05-0.3) | 12.3<br>(10.7-14.2) | 9.3<br>(8.6-10.1) | 6.8<br>(6.4 - 7.3) |
| Income | 24.8<br>(22.1-27.8) | 23.5<br>(21.4-25.8) | 26.0<br>(23.0-29.1) | 24.1<br>(21.9-26.4) | 20.4<br>(18.9-22.0) | 8.7<br>(5.4-13.6) | 36.8<br>(35.6-38.0) | 24.5<br>(22.9-26.1) | 13.1 (11.9-14.3) | 29.8 (27.5-32.2) | 27.48 (26.32-28.67) |  |
| Working hours/furlough | 39.8<br>(33.5-46.4) | 42.6<br>(39.9-45.4) | 26.9<br>(24.0-30.1) | 30.3<br>(28.3 - 32.4) | 41.2<br>(38.2 - 43.7) | 3.8<br>(2.6-5.5) | 51.8<br>(50.6-53.0) | 20.4<br>(18.9-22.0) | 6.5<br>(5.7-7.4) | 43.4<br>(40.5-46.5) | 11.0<br>(10.2 - 11.9) | 20.2<br>(19.4 - 20.9) |
| **Any housing disruption** | 35.8<br>(32.5-39.3) | 23.3<br>(21.6-25.2) | 15.2<br>(12.8-18.0) | 12.7<br>(11.3-14.1) | 9.7<br>(8.3-10.7) | 2.2<br>(1.3-3.7) | 31.8<br>(30.7-33.0) | 24.4<br>(22.8-26.1) | 7.7<br>(6.8-8.7) | 15.3<br>(13.9-16.8) | 6.8<br>(6.0-7.8) | 12.3<br>(11.8 - 12.9) |
| Housing loss/change | 2.4<br>(1.7-3.4) | 16.7<br>(15.2-18.4) | 1.4<br>(0.8-2.4) | 0.3<br>(0.2-0.6) | 0.01<br>(0.0-0.02) | 0 | 4.5<br>(4.0-5.1) | 1.8<br>(1.4-2.4) | 0.8<br>(0.5-1.2) | 2.1<br>(1.5-2.9) | 2.5<br>(2.1-2.9) | 3.8<br>(2.0 - 3.4) |
| Household composition | 35.4 (32.1-38.8) | 5.3 (4.4-6.4) | 15.1 (12.6-17.9) | 12.5 (11.2 - 14.0) | 9.7 (8.8 - 10.7) | 3.4<br>(2.0-5.6) | 29.2<br>(28.2-30.3) | 22.6<br>(21.1-24.2) | 7.7<br>(6.8-8.7) | 10.6<br>(9.5-11.8) | 4.5<br>(4.0-5.1) | 12.2<br>(11.6 - 12.7) |
| **Cumulative disruptions** |  |  |  |  |  |  |  |  |  |  |  |  |
| **No disruptions** | 35.1<br>(31.8-38.6) | 46.5<br>(44.5-48.6) | 47.2<br>(43.8-50.6) | 45.9<br>(43.4 - 48.4) | 49.2<br>(47.3-51.0) | 72.0<br>(66.7-76.8) | 23.0<br>(22.0-23.9) | 34.7<br>(33.1-36.4) | 52.4<br>(50.7-54.2) | 43.4<br>(41.3-45.5) | 60.8<br>(59-62.6) | 52.4<br>(51.5 - 53.3) |
| **Any one domain disrupted** | 43.0<br>(39.8-46.2) | 40.8<br>(38.8-42.8) | 39.0<br>(36.0-42.3) | 42.4<br>(40.0-44.8) | 40.5<br>(38.7-42.3) | 25.7<br>(21.0-31.0) | 43.9<br>(42.8-44.9) | 42.3<br>(40.5-44.1) | 39.7<br>(38.0-41.5) | 43.4<br>(41.3-45.6) | 31.5<br>(29.8-33.2) | 41.1<br>(40.2 - 41.9) |
| **Two domains disrupted** | 20.0<br>(17.4-22.8) | 11.7<br>(10.4-13.1) | 12.4<br>(10.1-15.0) | 10.8<br>(9.5-12.4) | 9.6<br>(8.6-10.7) | 2.3<br>(1.2-4.5) | 27.5<br>(26.5-28.4) | 20.0<br>(18.5-21.6) | 7.4<br>(6.5-8.3) | 11.7<br>(10.3-13.2) | 6.7<br>(5.8-7.7) | 6.5<br>(6.1 - 7.0) |
| **All three domains disrupted** | 1.9<br>(1.4-2.6) | 1.1<br>(0.7-1.6) | 1.4<br>(0.7-2.9) | 0.8<br>(0.6-1.2) | 0.7<br>(0.5-0.9) | 0 | 5.7<br>(5.2-6.3) | 3.0<br>(2.4-3.7) | 0.5<br>(0.2-0.8) | 1.5<br>(1.0-2.1) | 0.5<br>(0.3-0.8) | 0.02<br>(0.01 - 0.08) |

Sources: MCS (Millennium Cohort Study); ALSPAC G1 (Children of the Avon Longitudinal Study of Parents and Children); NS (Next Steps); BCS 70 (1970 British Cohort Study), NCDS (National Child Development Study); NSHD (National Survey of Health and Development); USoc (Understanding Society); ELSA (English Longitudinal Study of Ageing); GS (Generation Scotland: the Scottish Family Health Study); TWINSUK (UK Adult Twin Registry); COPING (Covid-19 Psychiatry and Neurological Genetics study), ALSPAC G0 (parents of ALSPAC). Weighted data. The detailed prevalence for each disruption can be found in the Supplementary Online file (see Supplementary Table S5). Note. TWINSUK had an additional question: “Have you experienced healthcare disruption as a result of the COVID-19 pandemic?” This data was also used to derive the ‘any healthcare disruption’ variable for TWINSUK.

### Pre-pandemic psychological distress and disruptions during the pandemic

The associations between standardised psychological distress and each outcome are illustrated in Figure 1. Table 3 shows the meta-analysed estimates for each outcome from the unadjusted, adjustment 1, and adjustment 2 models, and the heterogeneity in estimates (details of coefficients from each cohort and their weight in the meta-analysis for each outcome are available in supplementary file 3). Heterogeneity was lower in meta-analyses with greater adjustment and ranged from 0% to 66·8% across the different outcomes examined for the fully adjusted estimates.

**Figure 1.**
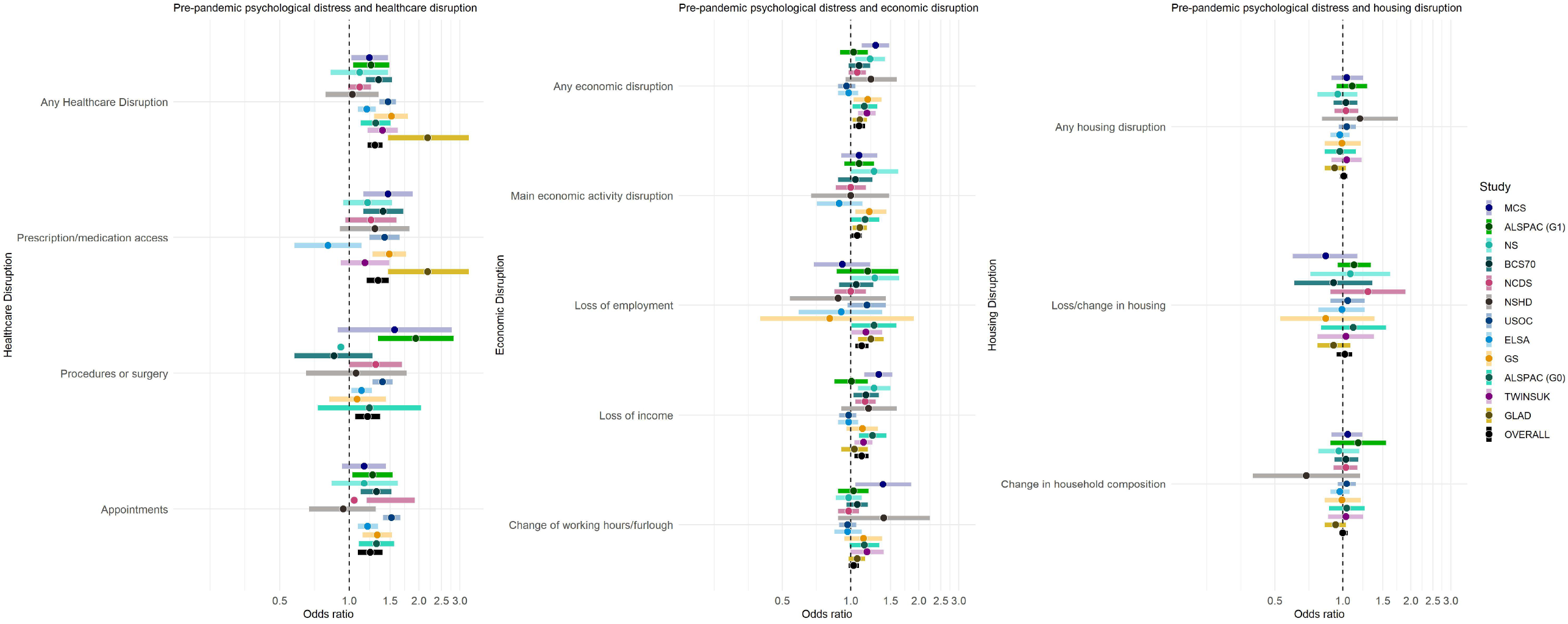
Odds Ratios between standardised psychological distress and each examined disruption. Estimates are adjusted for age, sex, ethnicity, education, UK Nation, partnership status, presence of children, housing tenure, occupational class, prior chronic conditions, and physical disability as appropriate and available in each study.

**Table 3.**
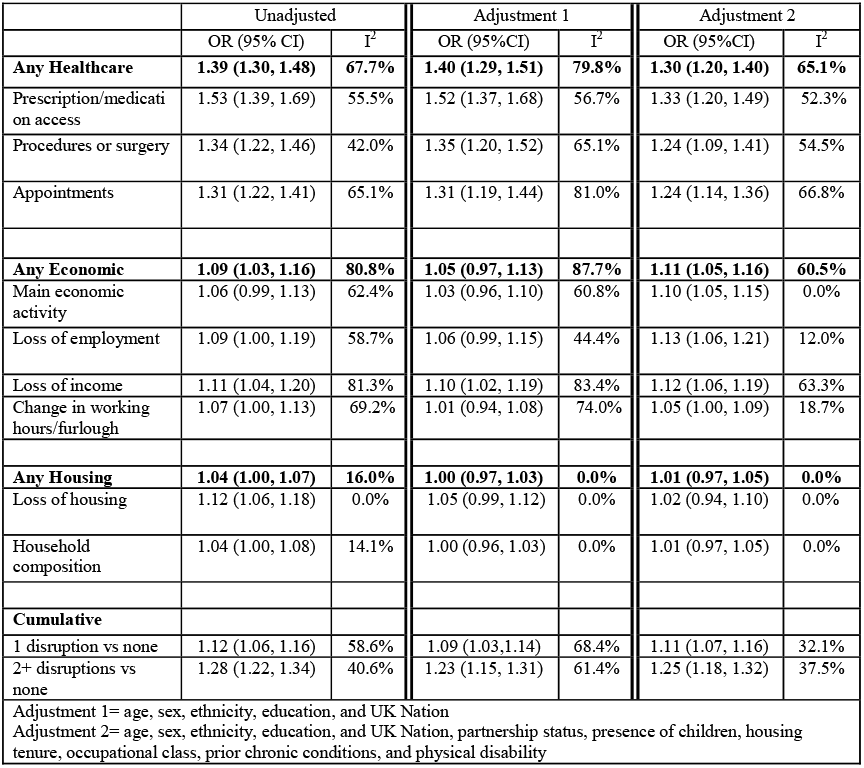
Meta-analysed associations between standardised psychological distress and healthcare, economic and housing disruptions.

|  | Unadjusted |  | Adjustment 1 |  | Adjustment 2 |  |
| --- | --- | --- | --- | --- | --- | --- |
|  | OR (95% CI) | I<sup>2</sup> | OR (95% CI) | I<sup>2</sup> | OR (95% CI) | I<sup>2</sup> |
| **Any Healthcare** | **1.39 (1.30, 1.48)** | **67.7%** | **1.40 (1.29, 1.51)** | **79.8%** | **1.30 (1.20, 1.40)** | **65.1%** |
| Prescription/medication access | 1.53 (1.39, 1.69) | 55.5% | 1.52 (1.37, 1.68) | 56.7% | 1.33 (1.20, 1.49) | 52.3% |
| Procedures or surgery | 1.34 (1.22, 1.46) | 42.0% | 1.35 (1.20, 1.52) | 65.1% | 1.24 (1.09, 1.41) | 54.5% |
| Appointments | 1.31 (1.22, 1.41) | 65.1% | 1.31 (1.19, 1.44) | 81.0% | 1.24 (1.14, 1.36) | 66.8% |
| **Any Economic** | **1.09 (1.03, 1.16)** | **80.8%** | **1.05 (0.97, 1.13)** | **87.7%** | **1.11 (1.05, 1.16)** | **60.5%** |
| Main economic activity | 1.06 (0.99, 1.13) | 62.4% | 1.03 (0.96, 1.10) | 60.8% | 1.10 (1.05, 1.15) | 0.0% |
| Loss of employment | 1.09 (1.00, 1.19) | 58.7% | 1.06 (0.99, 1.15) | 44.4% | 1.13 (1.06, 1.21) | 12.0% |
| Loss of income | 1.11 (1.04, 1.20) | 81.3% | 1.10 (1.02, 1.19) | 83.4% | 1.12 (1.06, 1.19) | 63.3% |
| Change in working hours/furlough | 1.07 (1.00, 1.13) | 69.2% | 1.01 (0.94, 1.08) | 74.0% | 1.05 (1.00, 1.09) | 18.7% |
| **Any Housing** | **1.04 (1.00, 1.07)** | **16.0%** | **1.00 (0.97, 1.03)** | **0.0%** | **1.01 (0.97, 1.05)** | **0.0%** |
| Loss of housing | 1.12 (1.06, 1.18) | 0.0% | 1.05 (0.99, 1.12) | 0.0% | 1.02 (0.94, 1.10) | 0.0% |
| Household composition | 1.04 (1.00, 1.08) | 14.1% | 1.00 (0.96, 1.03) | 0.0% | 1.01 (0.97, 1.05) | 0.0% |
| **Cumulative** |  |  |  |  |  |  |
| 1 disruption vs none | 1.12 (1.06, 1.16) | 58.6% | 1.09 (1.03, 1.14) | 68.4% | 1.11 (1.07, 1.16) | 32.1% |
| 2+ disruptions vs none | 1.28 (1.22, 1.34) | 40.6% | 1.23 (1.15, 1.31) | 61.4% | 1.25 (1.18, 1.32) | 37.5% |
| Adjustment 1= age, sex, ethnicity, education, and UK Nation |  |  |  |  |  |  |
| Adjustment 2= age, sex, ethnicity, education, and UK Nation, partnership status, presence of children, housing tenure, occupational class, prior chronic conditions, and physical disability |  |  |  |  |  |  |

In the fully adjusted models, one standard deviation higher psychological distress was associated with increased odds of any healthcare disruptions (OR 1·30 [95% CI: 1·20-1·40]) with ORs ranging from 1·24 to 1·33 for the different healthcare outcomes examined. ORs for each study were consistently >1 for all outcomes with a few exceptions, however, a substantial range was observed. For instance, ORs were between 1·03 and 1·53 for any healthcare disruption for the population representative cohorts, but higher (OR=2.18) in GLAD, which is a convenience sample with a higher proportion of participants with prior mental health difficulties.

For economic disruptions overall, one standard deviation higher psychological distress was associated with a higher likelihood of experiencing any economic disruption (OR=1·11 [95% CI: 1·05-1·16]), with associations found for loss of employment (OR= 1·13 [95% CI: 1·06-1·21]) and income (OR=1·12 [95% CI: 1·06-1·19]), and a smaller effect for reductions in working hours or furlough (OR= 1·05 [95% CI: 1·00 -1·09]). Some differences in study-level estimates were observed here, which likely reflect differences in study members’ ages. For instance, there were no observed associations with employment loss in older studies such as ELSA and NSHD, perhaps reflecting the lower proportions working post retirement age and the likelihood of those with good mental health being in this group.

There was no consistent evidence that prior psychological distress was associated with housing disruptions (OR= 1·01 [95% CI: 0·97, 1·05]).

1SD greater psychological distress prior to the pandemic was associated with an increased likelihood of experiencing disruption in at least two domains (RRR=1·25; [95% CI: 1·18, 1·32]) or in one domain (RRR=1·11; [95% CI: 1·07, 1·16]) relative to experiencing no disruption (Figure 2).

**Figure 2.**
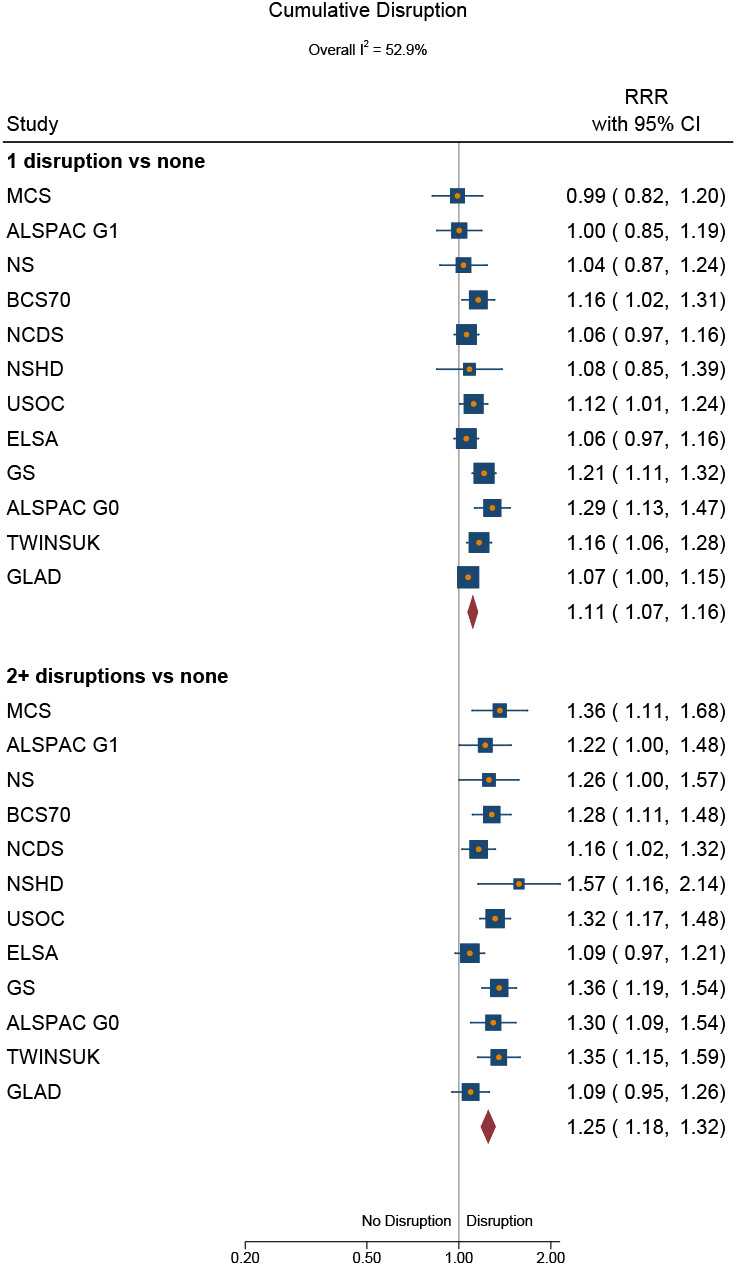
Associations between standardised psychological distress and cumulative disruptions. Models adjusted for age, sex, ethnicity, education, and UK Nation, partnership status, presence of children, housing tenure, occupational class, prior chronic conditions, and physical disability as appropriate and available in each study. RRR: Relative risk ratio.

Results from the meta-regression suggest that time since the pre-pandemic mental health measure does not explain the between-study heterogeneity (Supplementary Table S6).

### Stratified analyses

We explored subgroup differences in the associations between prior mental health and overall disruptions and found no evidence that associations differed by sex, education level, age, or ethnicity (see Supplementary Table S7).

### High psychological distress (binary indicator of caseness) as exposure

We conducted an additional analysis using a binary indicator of pre-pandemic high psychological distress. This was based on measure-specific cut-off scores which indicate clinical levels of distress (see results in supplementary file 3). Overall, findings were similar to those seen with continuous measures, with the largest associations seen for healthcare disruptions followed by economic disruptions and no associations for housing disruptions. However, owing to the different distribution and meaning of the dichotomised exposure, the observed effect sizes vary. For instance, based on this binary exposure, high psychological distress was associated with an increased likelihood of experiencing disruptions in at least two domains OR 1·46 (1·28, 1·67) compared to OR 1·18 (1·04, 1·33) in one domain relative to experiencing no disruption.

## Discussion

In our co-ordinated analysis of data from 12 UK-based longitudinal cohort studies, we found people with poor pre-pandemic mental health have experienced greater disruption to their lives across multiple domains during the COVID-19 pandemic. More specifically, (i) prior mental health difficulties were associated with greater likelihood of all examined healthcare disruptions (24-33% greater odds), economic disruptions (5-13% greater odds), and not associated with housing disruptions, (ii) the impact of prior mental health on these outcomes was not different by sex, education, age, or ethnicity, though pre-pandemic psychological distress was generally more common among women, younger generations, ethnic minorities, and those with fewer qualifications, and (iii) greater prior mental health difficulties were associated with greater likelihood of disruptions in multiple domains with 11% greater odds of disruption in one domain, and 25% greater odds of disruptions in two or three domains.

Healthcare disruptions have been widespread in the UK with numbers of treatments for non-COVID-19 illness dropping by millions compared to previous years (26). There has been a substantial decrease in the number of people attending A&E services (27), and reports of difficulties and delays accessing medication (28, 29). Reporting of healthcare disruptions ranged from under 10% to 37% across the included studies; this wide range may reflect both true gradients by age, and differences in sampling and assessment measures used (30). Disruptions associated with prior psychological distress included around a 24% greater odds of missed appointments and procedures and 33% greater odds of interruptions to prescriptions or medication access. Information on reasons for disruptions to healthcare access was not consistently available across studies, and could include: attempts to protect the NHS, patients or providers cancelling appointments, individuals being unable to rebook appointments, or being faced with complexities in the requirements for rebooking appointments or changing healthcare needs. Disruptions to healthcare are problematic both due to their potential longer-term adverse impacts on health outcomes and potential stress involved. Socio-demographic inequalities in healthcare access during the pandemic have been recorded across different data sources. Women, ethnic minorities, and those living in more deprived areas were more likely to experience healthcare disruptions (31, 32) and prior mental health might help explain some of these observed socio-demographic inequalities. Furthermore, since women, ethnic minorities, and those with lower levels of education were more likely to have experienced psychological distress before the pandemic, these mental health related disruptions to healthcare may also widen pre-pandemic social inequalities in health.

The pandemic has also impacted economic activity, with large numbers of people losing jobs, being put on furlough, and experiencing drops in household incomes (33, 34). Around 20-60% of individuals in working age cohorts reported disruptions to economic activity. As expected, this was lower in retired cohorts (e.g., 10% in the NSHD cohort who are now 75 years old). 1 SD of greater pre-pandemic psychological distress increased the likelihood of disruption by 10% to main economic activity, 13% to loss of employment, 12% to loss of income and 5% reductions in working hours or furlough. We did not examine potential positive economic outcomes such as starting a new business or increases in income or working hours. It is possible that there are differences in the ability of those with mental health difficulties to have economically benefitted or coped with additional or changed work demands during the pandemic. Younger workers, ethnic minorities, and females have been more likely to be in disrupted sectors and become unemployed or furloughed (35). Younger workers have been more likely to lose their jobs and report drops in income than older workers, reflecting their already more precarious labour market situation (34). However, the associations between prior mental health and poorer economic outcomes were not different across age and other socio-demographic groups. Again, given the socio-demographic inequalities in pre-pandemic mental health, this highlights how the pandemic may widen existing mental health and socio-demographic inequalities.

With overcrowded housing increasing risk of COVID-19 transmission, disparities in housing disruption are likely to have impacts on risk of COVID-19 infection and other poor health and economic outcomes (36). Although there have been reported changes in individuals’ housing situations during the pandemic, with evidence of younger people moving themselves, and older adults having people move into their households (37), we find no associations in the risk of housing disruptions with prior mental health in this study. This finding might reflect policies that were designed to minimise home loss during the pandemic and these outcomes should be monitored in the medium and longer-term as consequences of the economic and health disruptions are realised and protective policies are lifted. It is also plausible that participants who experienced adverse housing disruptions (e.g., homelessness) were less likely to participate in the COVID-19 surveys and might not be represented in these findings.

Across the included cohorts, around 25-45% of individuals reported at least one kind of disruption, with a further 2-30% across cohorts experiencing two out of three, and a smaller proportion (0.2-6.5%) experiencing all three. The heightened risk for clusters of disruptions for those with psychological distress may be largely due to the increased risk of disruptions to healthcare, economic activity and income, as that combined with no difference in risk for housing disruptions will still mean clusters of disruptions are more likely. Furthermore, adverse outcomes may cluster, for example, with housing disruption resulting from employment loss, or those with poor mental health being more likely to experience healthcare disruptions as a result of moving home and general practice (37). Multiple adverse disruptions are also potentially stressful and more predictive of poorer prognosis longer-term (38). We found that those with prior mental distress were more likely to suffer multiple disruptions, highlighting the need for inter-agency working in supporting those with mental ill-health.

## Strengths and Limitations

The analysis of multiple longitudinal cohorts with rich pre-COVID-19 information is an important strength of this study. Although many COVID-era online studies are available, the lack of pre-pandemic information makes it difficult to untangle the directions of associations between mental health and other outcomes. However, in the current study information on pre-pandemic healthcare use and disruptions were not consistently available across studies, so observed associations between pre-pandemic psychological distress and healthcare disruption may reflect being more likely to have healthcare needs to disrupt. This study is also strengthened by co-ordinated primary analysis in multiple longitudinal studies with differing study designs, different target populations, and varying selection and attrition processes. Heterogeneity in our meta-analysed estimates were often reduced when considering models with a greater number of possible confounders, highlighting the importance of adjusting for relevant pre-pandemic characteristics as appropriate for different generations and cohorts.

Differences between studies in a range of factors including measurement of mental health and outcomes, timing of surveys, design, response rates, and differential selection into the COVID-19 sweeps are potentially responsible for heterogeneity in estimates. However, despite this heterogeneity in the magnitude of estimates, the key findings were fairly consistent with regards to the direction of association across most studies. The differences might also be positively construed as allowing for replication and triangulation of findings that are robust to these intrinsic differences between studies. Furthermore, this heterogeneity can be informative, for example, by virtue of the mix of age-specific and age-range cohorts we could determine that the observed association between pre-pandemic psychological distress and disruptions does not differ by age.

## Implications and conclusions

Our findings highlight that people with poor mental health before the start of the pandemic were more likely to suffer negative economic and healthcare consequences in the first year of the pandemic, highlighting the need for policymakers to take this into account when provisioning current and post-pandemic health, economic, and well-being support. For instance, processes for re-booking healthcare procedures or accessing economic support should ensure that people struggling with mental health difficulties do not face additional barriers to accessing resources. Primary care practitioners and pharmacists should monitor patients with known mental health difficulties to ensure they do not miss appointments, procedures or prescriptions.

Individuals with mental health difficulties were more likely to have experienced adverse healthcare, economic, and housing outcomes even before the pandemic (7, 9). The pandemic created a situation where these disruptions were occurring at far greater rates than in a usual year. Given the far greater frequency of these disruptions in the population during COVID-19, the impacts on those with existing poor mental health will have been consequently larger.

Individuals with more severe mental disorders (e.g., schizophrenia, eating disorders), may have experienced even greater adversity from these disruptions, particularly in housing and economic domains. However, low prevalence of severe disorders generally leaves population-based samples underpowered to consider such conditions. Efforts to understand the impacts of the pandemic on those with more severe mental disorders is lacking, but needed. Current evidence suggests that they are at even greater risk of COVID-19 infection, mortality and non-vaccination uptake (5, 8).

Our findings highlight that many adverse socio-economic and health impacts of the pandemic have been disproportionately faced by those with prior mental ill-health, who are more likely to be women, those without a degree, and younger generations. The pandemic has the potential to increase social exclusion and widen existing physical health and economic inequalities amongst those with mental health problems, and mitigating this should be a public health priority. Ongoing monitoring is needed to get a full picture of the health and socio-economic implications of the pandemic for those with mental health difficulties.

## Data Availability

Data for NCDS (SN 6137), BCS70 (SN 8547), Next Steps (SN 5545), MCS (SN 8682) and all four COVID-19 surveys (SN 8658) are available through the UK Data Service. NSHD data are available on request to the NSHD Data Sharing Committee. Interested researchers can apply to access the NSHD data via a standard application procedure. Data requests should be submitted to; further details can be found at http://www.nshd.mrc.ac.uk/data.aspx. doi:10.5522/NSHD/Q101; doi:10.5522/NSHD/Q10.
ALSPAC data is available to researchers through an online proposal system. Information regarding access can be found on the ALSPAC website (http://www.bristol.ac.uk/media-library/sites/alspac/documents/researchers/data-access/ALSPAC_Access_Policy.pdf).
The TwinsUK Resource Executive Committee (TREC) oversees management, data sharing and collaborations involving the TwinsUK registry (for further details see https://twinsuk.ac.uk/resources-for-researchers/access-our-data/).
Understanding Society data are available through the UK Data Service (SN 6614 and SN 8644).
Waves 1-9 of ELSA are available through the UK Data Service (SN 8688 and 5050).
Access to Generation Scotland data is approved by the Generation Scotland Access Committee. See https://www.ed.ac.uk/generation-scotland/for-researchers/access or for further details.
Researchers wishing to access GLAD Study participants or data are invited to submit a data and sample access request to the NIHR BioResource to request a collaboration.

## Data sharing statement

All datasets included in this analysis have established data sharing processes, and for most included studies the anonymised datasets with corresponding documentation can be downloaded for use by researchers from the UK Data Service. We have detailed the exact processes for each dataset in Supplementary file 2.

## Acknowledgements

The contributing studies have been made possible because of the tireless dedication, commitment and enthusiasm of the many people who have taken part. We would like to thank the participants and the numerous team members involved in the studies including interviewers, technicians, researchers, administrators, managers, health professionals and volunteers. We are additionally grateful to our funders for their financial input and support in making this research happen.

GLAD: Alish Palmos, Christopher Hübel, Molly R Davies, Henry C Rogers, Yuhao Lin, Katherine S. Young, Thalia Eley, Matthew Hotopf, Kirstin Purves, Nathalie Kingston, John Bradley, Sofia Papadia, Debbie Clapham-Riley, Neil Walker.

GS: Drew Altschul, Chloe Fawns-Ritchie, Archie Campbell, Robin Flaig.

ALSPAC: Daniel J Smith, Nicholas J Timpson, Kate Northstone Understanding Society: Michaela Benzeval

NSHD: Andrew Wong, Maria Popham, Karen MacKinnon, Imran Shah, Philip Curran MCS, NS, BCS70, NCDS: Colleagues in survey, data and cohort maintenance teams

## Funding acknowledgements

This work was supported by the National Core Studies, an initiative funded by UKRI, NIHR and the Health and Safety Executive. The COVID-19 Longitudinal Health and Wellbeing National Core Study was funded by the Medical Research Council (MC_PC_20030).

## Studies

Understanding Society is an initiative funded by the Economic and Social Research Council and various Government Departments, with scientific leadership by the Institute for Social and Economic Research, University of Essex, and survey delivery by NatCen Social Research and Kantar Public. The Understanding Society COVID-19 study is funded by the Economic and Social Research Council (ES/K005146/1) and the Health Foundation (2076161). The research data are distributed by the UK Data Service.

The Millennium Cohort Study, Next Steps, 1970 British Cohort Study and 1958 National Child Development Study are supported by the Centre for Longitudinal Studies, Resource Centre 2015-20 grant (ES/M001660/1) and a host of other co-funders. The 1946 NSHD cohort is hosted by the the MRC Unit for Lifelong Health and Ageing funded by the Medical Research Council (MC_UU_00019/1Theme 1: Cohorts and Data Collection). The COVID-19 data collections in these five cohorts were funded by the UKRI grant Understanding the economic, social and health impacts of COVID-19 using lifetime data: evidence from 5 nationally representative UK cohorts (ES/V012789/1)

The English Longitudinal Study of Ageing was developed by a team of researchers based at University College London, NatCen Social Research, the Institute for Fiscal Studies, the University of Manchester and the University of East Anglia. The data were collected by NatCen Social Research. The funding is currently provided by the National Institute on Aging in the US, and a consortium of UK government departments coordinated by the National Institute for Health Research. Funding has also been received by the Economic and Social Research Council. The English Longitudinal Study of Ageing Covid-19 Substudy was supported by the UK Economic and Social Research Grant (ESRC) ES/V003941/1.

The UK Medical Research Council and Wellcome (Grant Ref: 217065/Z/19/Z) and the University of Bristol provide core support for ALSPAC. A comprehensive list of grants funding is available on the ALSPAC website (http://www.bristol.ac.uk/alspac/external/documents/grant-acknowledgements.pdf). We are extremely grateful to all the families who took part in this study, the midwives for their help in recruiting them, and the whole ALSPAC team, which includes interviewers, computer and laboratory technicians, clerical workers, research scientists, volunteers, managers, receptionists and nurses.

TwinsUK receives funding from the Wellcome Trust (WT212904/Z/18/Z), the National Institute for Health Research (NIHR) Biomedical Research Centre based at Guy’s and St Thomas’ NHS Foundation Trust and King’s College London. TwinsUK is also supported by the Chronic Disease Research Foundation and Zoe Global Ltd. The funders had no role in study design, data collection and analysis, decision to publish, or preparation of the manuscript.

Generation Scotland received core support from the Chief Scientist Office of the Scottish Government Health Directorates [CZD/16/6] and the Scottish Funding Council [HR03006]. Genotyping of the GS:SFHS samples was carried out by the Genetics Core Laboratory at the Wellcome Trust Clinical Research Facility, Edinburgh, Scotland and was funded by the Medical Research Council UK and the Wellcome Trust (Wellcome Trust Strategic Award “STratifying Resilience and Depression Longitudinally” (STRADL) Reference 104036/Z/14/Z). Generation Scotland is funded by the Wellcome Trust (216767/Z/19/Z).

The Genetic Links to Anxiety and Depression project is supported by the National Institute for Health Research (NIHR) BioResource, the NIHR BioResource Centre Maudsley, and the Biomedical Research Centre at South London and Maudsley NHS Foundation Trust and King’s College London. This study presents independent research supported by the National Institute for Health Research (NIHR) Biomedical Research Centre BioResource at South London and Maudsley NHS Foundation Trust and King’s College London. The views expressed are those of the author(s) and not necessarily those of the NHS, NIHR, Department of Health and Social Care or King’s College London. We gratefully acknowledge capital equipment funding from the Maudsley Charity (Grant Ref. 980) and Guy’s and St Thomas’s Charity (Grant Ref. STR130505).

SVK acknowledges funding from a NRS Senior Clinical Fellowship (SCAF/15/02), the Medical Research Council (MC_UU_00022/2) and the Scottish Government Chief Scientist Office (SPHSU13). ASFK acknowledges funding from the ESRC (ES/V011650/1). DJP acknowledges funding from the Wellcome Trust (216767/Z/19/Z and 221574/Z/20/Z). CLN acknowledges funding from a Medical Research Council Fellowship (MR/R024774/1). EJT acknowledges funding from the Wellcome Trust (WT212904/Z/18/Z).

Role of funder. The funders had no role in the methodology, analysis or interpretation of the findings presented in this manuscript.

## Author Contribution Statement

Patalay, Porteous, and Chaturvedi conceptualised the study and design. Patalay, Green, Thompson, Di Gessa, McElroy, Maddock, Katikireddi, Niedzwiedz, Griffith, Kwong, and Silverwood designed the methodology. Green, Thompson, Di Gessa, McElroy, Maddock, Stevenson, Davies, Mundy, Griffith, and Kwong conducted the formal analysis. Green, Thompson, Di Gessa, McElroy, Maddock, Stevenson, Griffith, Kwong, Steves, Chaturvedi, Henderson, and Fitzsimons were responsible for data curation. Patalay, Green, Di Gessa, and Maddock wrote the original draft of the manuscript. All authors contributed to critical revision of the manuscript. Thompson, Di Gessa, Maddock contributed to data visualisation. The project was supervised by Patalay, Porteous, and Katikireddi. Funding was acquired by Patalay, Katikireddi, Breen, Porteous, Steptoe, Ploubidis, Silverwood, Steves and Chaturvedi.

