## Supplementary file 2 for "Pre-pandemic mental health and disruptions to healthcare, economic, and housing outcomes during COVID –19: evidence from 12 UK longitudinal studies"

### Mental health inequalities in healthcare, economic, and housing disruption during COVID -19: an investigation in 12 longitudinal studies

### Supplementary file 2

### S2.1 Ethics and data access statements for each study

The most recent sweeps of the **NSHD, NCDS, BCS70**, **Next Steps** and **MCS** have all been granted ethical approval by the National Health Service (NHS) Research Ethics Committee and all participants have given informed consent. Data for NCDS (SN 6137), BCS70 (SN 8547), Next Steps (SN 5545), MCS (SN 8682) and all four COVID-19 surveys (SN 8658) are available through the UK Data Service. NSHD data are available on request to the NSHD Data Sharing Committee. Interested researchers can apply to access the NSHD data via a standard application procedure. Data requests should be submitted to; further details can be found at http://www.nshd.mrc.ac.uk/data.aspx. doi:10.5522/NSHD/Q101; doi:10.5522/NSHD/Q10.

Ethical approval was obtained from the **ALSPAC** Ethics and Law Committee and the Local Research Ethics Committees. The study website contains details of all the data that is available through a fully searchable data dictionary and variable search tool: http://www.bristol.ac.uk/alspac/researchers/our-data. ALSPAC data is available to researchers through an online proposal system. Information regarding access can be found on the ALSPAC website (http://www.bristol.ac.uk/media-library/sites/alspac/documents/researchers/data-access/ALSPAC_Access_Policy.pdf).

All wave of **TwinsUK** have received ethical approval associated with TwinsUK Biobank (19/NW/0187), TwinsUK (EC04/015) or Healthy Ageing Twin Study (H.A.T.S) (07/H0802/84) studies from NHS Research Ethics Committees at the Department of Twin Research and Genetic Epidemiology, King's College London. The TwinsUK Resource Executive Committee (TREC) oversees management, data sharing and collaborations involving the TwinsUK registry (for further details see https://twinsuk.ac.uk/resources-for-researchers/access-our-data/).

The University of Essex Ethics Committee has approved all data collection for the **Understanding Society** main study and COVID-19 waves. No additional ethical approval was necessary for this secondary data analysis. All data are available through the UK Data Service (SN 6614 and SN 8644).

Waves 1-9 of **ELSA** were approved through the National Research Ethics Service, while the COVID-19 Sub-study was approved by the UCL Research Ethics Committee. All participants provided informed consent. All data are available through the UK Data Service (SN 8688 and 5050).

**Generation Scotland** obtained ethical approval from the East of Scotland Committee on Medical Research Ethics (on behalf of the National Health Service). Reference number 20/ES/0021. Access to data is approved by the Generation Scotland Access Committee. See https://www.ed.ac.uk/generation-scotland/for-researchers/access or for further details.

The **GLAD** Study was approved by the London - Fulham Research Ethics Committee on 21st August 2018 (REC reference: 18/LO/1218) following a full review by the committee. Researchers wishing to access GLAD Study participants or data are invited to submit a data and sample access request to the NIHR BioResource to request a collaboration.

### S2.2 Further details of measures of psychological distress

**MCS:** The K-6[1] is a 6-item measure of psychological distress (i.e., general anxiety and depression). Responses are rated on a 5-point Likert-type scale, and capture distress over a period of four weeks prior to administration of the scale. Scores range from 0 to 24, with a conservative cut-off of 13+ applied to indicate probable psychological distress.

**NCDS, BCS70**: The 9-item version of the Malaise Inventory[2] was used to assess general psychological distress. Items are scored using a simple ‘Yes/No’ response, meaning continuous scores range from 0-9. Scores of four or more are indicative of probable psychiatric distress.

**Understanding Society**, **Next Steps, Generation Scotland, NSHD**: The 28-item General Health Questionnaire (GHQ)[3] was used to detect symptoms of psychological distress in GS and NSHD. The GHQ is a screening instrument designed to detect symptoms of psychological distress (i.e. general anxiety and depression). Each item is scored 0-3 resulting in scores ranging from 0-84. There is an alternative scoring where each item is scored as 0-0-1-1. The 12-item GHQ was used to detect symptoms of psychological distress in Understanding Society and Next Steps. This version is scored in the same manner as the 28-item questionnaire, resulting in scores ranging from 0-36.

**GLAD**: The Patient Health Questionnaire (PHQ-9)[4] is a 9-item tool used by healthcare professionals to assess severity of depressive symptoms. Individuals are asked to indicate, from 0 “Not at all” to 3 “Nearly every day” how often they have been bothered by problems such as “Little interest or pleasure in doing things?”. Each answer is scored between 0 and 3, leaving each participant with a total score out of 27.

**ALSPAC G1**: Self-reported depressive symptoms were measured using the short mood and feelings questionnaire (SMFQ).[5] The SMFQ is a 13-item questionnaire that measures the presence of depression symptoms in the previous two weeks and was administered via postal questionnaire or in research clinics. Each item is scored between 0-2, resulting in a summed score between 0-26. Depression severity can be rated in the following score bands: 0-4 none, 5-9 mild, 10-14 moderate, 15-19 moderately severe, 20-27 severe.

**ALSPAC G0**: The Edinburgh Postnatal Depression Scale (EPDS)[6] was originally developed to screen for postnatal depression in women, but has since been shown to effectively screen for depression in men also.[7] This 10-item questionnaire assesses the severity of depressive symptoms over the previous 7 days using a 4-point Likert response scale. Cut-off values of 13 or higher are most often used to identify those who might have probable depression.

**ELSA**: Depressive symptoms were measured using the 8-item version of the Center for Epidemiologic Studies Depression Scale (CES-D).[8] This measure asks respondents to indicate how often they experienced symptoms over the previous week using a 4-point Likert response. A binary (present/absent) scoring system was applied in the present study.

**TwinsUK**: The Hospital Anxiety and Depression Scale (HADS)[9] is a 14-item scale used to measure levels of psychiatric distress in non-psychiatric patient populations. Responses are indicated on a 4-point ordinal Likert scale.

#### S2.3 Supplementary Tables

##### Supplementary Table S1. Percent (and N) distribution of demographic and socio-economic characteristics by study

| | ***MCS*** | ***ALSPAC G1*** | ***NS*** | ***BCS 70*** | ***NCDS*** | ***NSHD*** | ***USOC*** | ***ELSA*** | ***GS*** | ***ALSPAC G0*** | ***TWINS UK*** | ***GLAD*** |
|---|---|---|---|---|---|---|---|---|---|---|---|---|
| **Total analytic N** | 3,028 | 2,698 | 3,209 | 4,303 | 5,394 | 1,310 | 13,175 | 5,061 | 3,179 | 3,212 | 2855 | 12,107 |
| Female | 65.5 (2,007) | 68.1 (1,837) | 64.7 (2,077) | 57.8 (2,494) | 53.6 (2,892) | 52.8 (691) | 57.9 (7,623) | 57.1 (2,891) | 63.7 (2,025) | 72.5 (2,327) | 89.8 (2,565) | 82.1 (9,935) |
| Mean age in 2020 (range) | 19.5 (18-20) | 27.6 (27-29) | 30.6 (29-31) | 50.5 | 62.6 | 74.0 | 51.2 (16-96) | 70.3 (52-90+) | 60.1 (27-100) | 58.8 (45-81) | 64.4 (22-96) | 43.0 (16-89) |
| Ethnicity | | | | | | | | | | | | |
| White | 86.1 (2,636) | 96.7 (2,608) | 75.1 (2,409) | -- | -- | -- | 87.4 (11,517) | 96.4 (4,881) | 99.3 (3,157) | 98.5 (3,143) | 98.5 (2,811) | 95.8 (11,572) |
| South Asian | 8.3 (255) | -- | 16.5 (528) | -- | -- | -- | 6.5 (861) | 1.8 (88) | 0.2 (7) | -- | 0.2 (6) | -- |
| Black | 2.7 (82) | -- | 3.9 (126) | -- | -- | -- | 2.5 (329) | 1.0 (50) | 0 (1) | -- | 0.6 (18) | 0.3 (37) |
| Mixed | 2.5 (76) | -- | 4.6 (146) | -- | -- | -- | 1.8 (238) | 0.8 (42) | 0.4 (13) | -- | 0.5 (15) | 2.1 (259) |
| Other | 0.5 (14) | -- | 0 | -- | -- | -- | 1.8 (230) | -- | 0 (1) | -- | 0.3 (5) | 0.9 (108) |
| Ethnic minority | 13.9 (427) | 3.3 (88) | 24.9 (800) | -- | -- | -- | 12.6 (1,658) | 3.6 (180) | 0.7 (22) | 1.5 (48) | 1.5 (44) | 4.2 (507) |
| High Education | 56.5 (1,729) | 20.4 (551) | 49.9 (1,600) | 47.6 (2,053) | 46.4 (2,504) | 29.9 (391) | 47.2 (6,219) | 25.6 (1,297) | 46.6 (1,483) | 30.2 (969) | 52.9 (1,509) | 59.0 (7,138) |
| Social class | | | | | | | | | | | | |
| Managerial, Admin, and Professional occupation | 60.5 (1,560) | 45.6 (1,066) | 54.0 (1,461) | 67.6 (2,797) | 60.9 (3,075) | 56.0 (733) | 35.1 (4,624) | 34.0 (1,720) | 61.0 (1,939) | 52.3 (1,520) | -- | -- |
| Intermediate occupations | 17.9 (461) | 53.2 (1,246) | 23.5 (635) | 18.4 (763) | 21.8 (1,098) | 34.5 (452) | 17.1 (2,250) | 24.1 (1,221) | 11.2 (355) | 47.1 (1,369) | -- | -- |
| Routine and Manual occupations | 21.6 (556) | 1.2 (28) | 22.5 (608) | 13.7 (567) | 16.8 (846) | 9.2 (121) | 20.1 (2,641) | 28.4 (1,438) | 3.3 (104) | 0.6 (17) | -- | -- |
| Never worked, long-term unemployed, or missing | -- | -- | -- | 0.3 (11) | 0.5 (27) | 0.3 (4) | 27.8 (3,660) | 13.5 (682) | 24.5 (781) | -- | -- | -- |
| Country of residence | | | | | | | | | | | | |
| England | 67.5 (2,066) | -- | 96.8 (3,105) | 86.0 (3,709) | 84.1 (4,536) | 87.0 (1,140) | 84.1 (11,078) | 100 (5,061) | 16 (0.5) | -- | 92.5 (2645) | 89.5 (10,837) |
| Scotland | 13.0 (397) | -- | 0.6 (20) | 8.1 (351) | 8.8 (476) | 7.6 (99) | 7.5 (983) | -- | 99.4 (3,160) | -- | 3.6 (104) | 5.4 (654) |

| | | | | | | | | | | | | |
|---|---|---|---|---|---|---|---|---|---|---|---|---|
| Wales | 11.5 (351) | -- | 0.6 (19) | 5.0 (215) | 5.2 (281) | 4.3 (56) | 5.1 (669) | -- | -- | -- | 3.2 (92) | 2.5 (307) |
| N.Ireland | 7.4 (227) | -- | 0.2 (5) | 0.1 (2) | 0.1 (3) | -- | 3.4 (445) | -- | -- | -- | 0.5 (14) | 2.6 (309) |
| Other | 0.7 (22) | -- | 1.9 (60) | 0.8 (35) | 1.8 (98) | 0.7 (9) | -- | -- | 0.1 (3) | -- | -- | -- |

Sources: MCS (Millennium Cohort Study); ALSPAC G1 (Children of the Avon Longitudinal Study of Parents and Children); NS (Next Steps); BCS 70 (1970 British Cohort Study), NCDS (National Child Development Study); NSHD (National Survey of Health and Development); USOC (Understanding Society); ELSA (English Longitudinal Study of Ageing); GS (Generation Scotland: the Scottish Family Health Study); TWINS UK (UK Adult Twin Registry); GLAD (Genetic Links to Anxiety and Depression), ALSPAC G0 (parents of ALSPAC). Notes: Samples for each study restricted to respondents with non-missing pre-pandemic psychological distress measure, with at least one disruption experienced during the pandemic, and valid information on sex and age. -- (not available/applicable).

**Supplementary Table S2. Mean pre-pandemic psychological distress scores and % with high psychological distress, by study**

| | ***MCS*** | ***ALSPAC G1*** | ***NS*** | ***BCS70*** | ***NCDS*** | ***NSHD*** | ***USOC*** | ***ELSA*** | ***GS*** | ***ALSPAC G0*** | ***TWINS UK*** | ***GLAD*** |
|---|---|---|---|---|---|---|---|---|---|---|---|---|
| **Measure** | K6 | SMFQ | GHQ-12 | Malaise | Malaise | GHQ-28 | GHQ-12 | CES-D | GHQ-28 | EPDS | HADS | PHQ9 |
| **Range** | 0-24 | 0-26 | 0-36 | 0-9 | 0-9 | 0-27 | 0-36 | 0-8 | 0-70 | 0-30 | 0-36 | 0-27 |
| **Mean (SD)** | 7.7 (5.0) | 6.7 (6.2) | 11.9 (6.0) | 1.9 (2.2) | 1.4 (1.9) | 1.7 (3.3) | 11.5 (8.4) | 1.4 (1.9) | 14.9 (7.7) | 6.4 (5.5) | 7.4 (6.0) | 11.2 (6.9) |
| **% High psychological distress** | 18.4 | 23.9 | 24.3 | 21.8 | 13.2 | 15.2 | 20.1 | 12.8 | 11.8 | 18.3 | 5.8 | 54.5 |
| **Threshold for high psychological distress** | 13+ | 11+ | 4+ | 4+ | 4+ | 4+ | 4+ | 4+ | 24+ | 12+ | 11+ | 10+ |
| **Year of pre-pandemic assessment** | 2018 | 2017/18 | 2015 | 2016 | 2008 | 2015 | 2018/19 | 2018/19 | 2006/11 | 2011/13 | 2017/18 | 2018/20 |
| **Gap (in years) to pandemic** | 2 | 2/3 | 5 | 4 | 12 | 5 | 1/2 | 1/2 | 9/14 | 7/9 | 2/3 | 0/2 |
| **Mean age when assessed** | 17.3 | 25.3 | 26.0 | 46.9 | 50.7 | 69.0 | 48.0 | 68.4 | 49.6 | 51.1 | 65.3 | 42.1 |

Sources: MCS (Millennium Cohort Study); ALSPAC G1 (Children of the Avon Longitudinal Study of Parents and Children); NS (Next Steps); BCS 70 (1970 British Cohort Study), NCDS (National Child Development Study); NSHD (National Survey of Health and Development); USOC (Understanding Society); ELSA (English Longitudinal Study of Ageing); GS (Generation Scotland: the Scottish Family Health Study); TWINS UK (UK Adult Twin Registry); GLAD (Genetic Links to Anxiety and Depression), ALSPAC G0 (parents of ALSPAC). Weighted data.

**Supplementary Table S3. Mean pre-pandemic psychological distress scores (and 95% confidence intervals) by socio-demographic characteristics and study**

| | *MCS* | *ALSPAC G1* | *NS* | *BCS 70* | *NCDS* | *NSHD* | *USOC* | *ELSA* | *GS* | *ALSPAC G0* | *TWINS UK* | *GLAD* |
|---|---|---|---|---|---|---|---|---|---|---|---|---|
| **Male** | 6.8 (6.3-7.2) | 5.3 (4.9-5.7) | 11.4 (11.0-11.7) | 1.6 (1.4-1.7) | 1.1 (1.0-1.2) | 1.0 (0.7-1.3) | 10.8 (10.6-11.0) | 1.1 (1.0-1.25) | 13.9 (13.5-14.3) | 5.4 (5.0-5.8) | 6.2 (5.6-6.8) | 10.8 (10.5-11.1) |
| **Female** | 8.7 (8.2-9.1) | 7.4 (7.1-7.7) | 12.4 (12.1-12.6) | 2.3 (2.1-2.4) | 1.67 (1.6-1.8) | 2.3 (1.7-3.0) | 12.1 (11.9-12.3) | 1.7 (1.6-1.8) | 15.6 (15.2-15.9) | 6.8 (6.5-7.1) | 7.6 (7.3-7.8) | 11.3 (11.1-11.4) |
| **No high education** | 8.0 (7.5-8.6) | 6.9 (6.6-7.2) | 12.3 (11.9-12.6) | 2.1 (1.9-2.3) | 1.51 (1.4-1.6) | 1.9 (1.4-2.4) | 11.6 (11.4-11.8) | 1.5 (1.4-1.6) | 15.1 (14.7-15.5) | 6.2 (6.3-6.9) | 7.6 (7.3-8.0) | 12.7 (12.5-12.8) |
| **High education** | 7.4 (7.0-7.7) | 6.2 (5.6-6.7) | 11.8 (11.5-12.1) | 1.5 (1.4-1.6) | 1.22 (1.2-1.3) | 1.1 (0.6-1.5) | 11.3 (11.1-11.5) | 1.1 (0.9-1.2) | 14.8 (14.4-15.2) | 5.8 (5.5-6.2) | 7.2 (6.9-7.5) | 10.2 (10.0-10.3) |
| **White** | 7.8 (7.5-8.2) | 6.7 (6.4-7.0) | 12.1 (11.9-12.4) | -- | -- | -- | 11.5 (11.3-11.6) | 1.4 (1.3-1.4) | 14.9 (14.7-15.2) | 6.5 (6.3-6.7) | 7.4 (7.2-7.6) | 11.1 (11.0-11.2) |
| **Ethnic minority** | 7.28 (6.8-7.7) | 7.8 (5.8-9.9) | 11.8 (11.3-12.2) | -- | -- | -- | 11.9 (11.3-12.4) | 2.4 (1.8-2.9) | 17.5 (12.4-22.7) | 5.6 (4.1-7.1) | 9.1 (6.9-11.3) | 12.4 (11.7-13.1) |
| **16-24** | -- | -- | -- | -- | -- | -- | 12.4 (12.0-12.9) | -- | -- | -- | 6.9 (-2.1-15.8) | 14.0 (13.6-14.3) |
| **25-34** | -- | -- | -- | -- | -- | -- | 12.1 (11.6-12.6) | -- | 15.6 (13.7-17.5) | -- | 9.5 (8.3-11) | 11.6 (11.3-11.8) |
| **35-44** | -- | -- | -- | -- | -- | -- | 12.0 (11.6-12.3) | -- | 15.4 (14.5-16.3) | -- | 8.5 (7.5-9.4) | 11.0 (10.7-11.3) |
| **45-54** | -- | -- | -- | -- | -- | -- | 11.9 (11.6-12.2) | 1.4 (1.1-1.6) | 15.3 (14.6-15.9) | 7.1 (6.2-8.1) | 8.1 (7.4-8.9) | 11.2 (10.9-11.4) |
| **55-64** | -- | -- | -- | -- | -- | -- | 11.5 (11.2-11.8) | 1.5 (1.4-1.7) | 16.1 (15.6-16.6) | 6.4 (6.1-6.7) | 7.9 (7.4-8.4) | 10.1 (9.8-10.4) |
| **65-74** | -- | -- | -- | -- | -- | -- | 10.0 (9.8-10.3) | 1.3 (1.2-1.4) | 14.1 (13.7-14.5) | 5.5 (4.8-6.3) | 6.8 (6.5-7.2) | 7.8 (7.4 - 8.2) |

| | | | | | | | | | | | | |
|---|---|---|---|---|---|---|---|---|---|---|---|---|
| **75+** | -- | -- | -- | -- | -- | -- | 9.9 (9.6-10.3) | 1.5 (1.4-1.6) | 12.4 (11.5-13.3) | 5.6 (2.2-8.9) | 7.0 (6.6-7.4) | 6.9 (5.8-8.1) |

Sources: MCS (Millennium Cohort Study); ALSPAC G1 (Children of the Avon Longitudinal Study of Parents and Children); NS (Next Steps); BCS 70 (1970 British Cohort Study), NCDS (National Child Development Study); NSHD (National Survey of Health and Development); USOC (Understanding Society); ELSA (English Longitudinal Study of Ageing); GS (Generation Scotland: the Scottish Family Health Study); TWINS UK (UK Adult Twin Registry); GLAD (Genetic Links to Anxiety and Depression), ALSPAC G0 (parents of ALSPAC). Weighted data.

Notes: -- (not available/applicable).

**Supplementary Table S4. Percentage with high psychological distress scores (and 95% confidence intervals) by socio-demographic characteristics and study**

| | *MCS* | *ALSPAC G1* | *NS* | *BCS 70* | *NCDS* | *NSHD* | *USOC* | *ELSA* | *GS* | *ALSPAC G0* | *TWINS UK* | *GLAD* |
|---|---|---|---|---|---|---|---|---|---|---|---|---|
| **Male** | 13.3 (9.9-17.7) | 16.7 (13.7-20.0) | 22.7 (20.4-25.2) | 17.5 (14.3-21.2) | 9.6 (7.9-11.6) | 7.1 (4.3-11.6) | 16.4 (15.2-17.8) | 9.1 (7.4-11.1) | 9.4 (7.7-11.2) | 10.7 (7.7-14.7) | 2.4 (1.2-5.0) | 51.43 (49.3-53.5) |
| **Female** | 23.5 (20.3-27.1) | 27.2 (24.6-30.1) | 28.1 (26.2-30.0) | 26.1 (23.0-30.0) | 16.7 (15.0-18.6) | 22.1 (15.3-30.8) | 23.5 (22.2-24.8) | 16.1 (14.2-18.0) | 13.1 (11.7-14.7) | 20.9 (18.7-23.4) | 6.2 (5.3-7.2) | 55.18 (55.0-56.2) |
| **No high education** | 22.5 (18.1-27.7) | 25.2 (22.8-27.8) | 27.2 (25.0-29.4) | 25.3 (22.0-29.0) | 14.6 (12.8-16.5) | 16.0 (11.2-22.2) | 20.6 (19.3-21.9) | 14.1 (12.6-15.8) | 12.9 (11.3-14.6) | 19.4 (17.1-21.9) | 5.6 (4.5-7.0) | 63.41 (62.1-64.7) |
| **High education** | 14.4 (11.8-17.3) | 19.0 (15.1-23.6) | 25.2 (23.1-27.4) | 15.3 (13.4-17.3) | 10.9 (9.5-12.5) | 11.7 (5.6-22.7) | 19.4 (18.1-20.7) | 8.5 (6.5-10.9) | 10.4 (8.9-12.1) | 14.3 (11.9-16.9) | 5.9 (4.8-7.2) | 48.30 (47.2-49.5) |
| **White** | 19.2 (16.3-22.3) | 23.7 (21.5-25.9) | 25.7 (23.9-27.4) | -- | -- | -- | 19.7 (18.7-20.7) | 11.8 (10.6-13.0) | 11.6 (10.5-12.8) | 18.4 (16.4-20.5) | 5.7 (4.9-6.6) | 54.20 (53.3-55.1) |
| **Ethnic minority** | 15.7 (12.5-19.5) | 32.7 (19.9-48.7) | 27.8 (24.8-31.0) | -- | -- | -- | 24.8 (21.3-28.6) | 27.1 (18.6-37.7) | 22.7 (7.8-45.4) | 13.0 (5.0-29.8) | 9.1 (4.4-22.1) | 60.95 (56.6-65.1) |
| **16-24** | -- | -- | -- | -- | -- | -- | 29.9 (26.7-33.4) | -- | -- | -- | 12.5 (1.5-57.3) | 73.2 (71.0-75.4) |
| **25-34** | -- | -- | -- | -- | -- | -- | 22.3 (19.3-25.5) | -- | 16.0 (9.6-24.4) | -- | 14.6 (8.3-24.7) | 58.3 (56.4-60.1) |
| **35-44** | -- | -- | -- | -- | -- | -- | 22.5 (20.1-25.0) | -- | 12.6 (8.7-17.3) | -- | 8.4 (5.2-13.2) | 53.5 (51.5-55.5) |
| **45-54** | -- | -- | -- | -- | -- | -- | 20.9 (18.9-23.0) | 12.1 (8.7-16.6) | 12.4 (9.7-15.4) | 23.7 (17.7-31.0) | 9.2 (6.4-13.1) | 52.8 (50.9-54.6) |

| | | | | | | | | | | | | |
|---|---|---|---|---|---|---|---|---|---|---|---|---|
| **55-64** | -- | -- | -- | -- | -- | -- | 19.1 (17.3-21.0) | 15.1 (12.3-18.3) | 14.2 (12.0-16.7) | 17.5 (15.6-19.6) | 6.5 (4.8-8.7) | 47.1 (44.9-49.4) |
| **65-74** | -- | -- | -- | -- | -- | -- | 11.7 (10.2-13.5) | 10.9 (9.4-12.6) | 9.8 (8.3-11.7) | 14.0 (1.0-19.5) | 4.8 (3.7-6.2) | 32.9 (29.6-36.4) |
| **75+** | -- | -- | -- | -- | -- | -- | 11.8 (9.5-14.4) | 12.3 (9.4-12.6) | 5.5 (2.4-10.5) | <0.1 | 3.0 (1.8-4.7) | 30.2 (21.3-40.9) |

Sources: MCS (Millennium Cohort Study); ALSPAC G1 (Children of the Avon Longitudinal Study of Parents and Children); NS (Next Steps); BCS 70 (1970 British Cohort Study), NCDS (National Child Development Study); NSHD (National Survey of Health and Development); USOC (Understanding Society); ELSA (English Longitudinal Study of Ageing); GS (Generation Scotland: the Scottish Family Health Study); TWINS UK (UK Adult Twin Registry); GLAD (Genetic Links to Anxiety and Depression), ALSPAC G0 (parents of ALSPAC). Weighted data.

Notes: -- (not available/applicable).

**Supplementary Table S5. Percent prevalence of any healthcare, economic, and housing disruptions during the pandemic by socio-demographic characteristics and study**

| | *Sex* | | *Ethnicity* | | *Education* | | *Age group* | | | | | | |
|---|---|---|---|---|---|---|---|---|---|---|---|---|---|
| | **Male** | **Female** | **White** | **Ethnic minority** | **Not high** | **High** | **16-24** | **25-34** | **35-44** | **45-54** | **55-64** | **65-74** | **75+** |
| **Any healthcare disruption** | | | | | | | | | | | | | |
| **MCS** | 6.6 | 12.4 | 10.6 | 9.6 | 9.8 | 10.9 | 10.4 | -- | -- | -- | -- | -- | -- |
| **ALSPAC-G1** | 12.8 | 17.5 | 16.1 | 9.0 | 15.8 | 16.4 | -- | 15.9 | -- | -- | -- | -- | -- |
| **NS** | 8.0 | 12.7 | 11.5 | 9.3 | 10.7 | 11.3 | -- | 11.0 | -- | -- | -- | -- | -- |
| **BCS 70** | 10.0 | 15.6 | -- | -- | 13.3 | 13.2 | -- | -- | -- | 13.2 | -- | -- | -- |
| **NCDS** | 13.9 | 15.2 | -- | -- | 14.2 | 15.1 | -- | -- | -- | -- | 14.6 | -- | -- |
| **NSHD** | 17.3 | 19.3 | -- | -- | 18.3 | 18.4 | -- | -- | -- | -- | -- | 18.3 | -- |
| **USOC** | 29.4 | 34.1 | 32.0 | 30.0 | 33.2 | 29.6 | 18.5 | 24.4 | 24.9 | 30.8 | 38.5 | 43.5 | 45.3 |
| **ELSA** | 36.4 | 36.9 | 36.4 | 40.4 | 37.1 | 36.0 | -- | -- | -- | 32.6 | 35.8 | 36.2 | 40.5 |
| **GS** | 27.4 | 27.5 | 27.4 | 36.4 | 29 | 25.6 | -- | 24.5 | 22.4 | 24.1 | 25 | 30.6 | 39 |
| **ALSPAC-G0** | 18.1 | 20.5 | 19.9 | 25.6 | 20.1 | 19.4 | -- | -- | -- | 21.4 | 19.2 | 21.6 | 30.6 |
| **TWINSUK** | 10.3 | 8.5 | 8.7 | 9.1 | 8.0 | 9.3 | 12.5 | 8.0 | 14.7 | 9.6 | 8.1 | 9.2 | 6.0 |
| **GLAD** | 0.8 | 0.7 | 0.7 | 0.5 | 0.7 | 0.7 | 0.7 | 0.9 | 0.8 | 0.5 | 0.6 | 0.8 | 1.3 |
| **Any economic disruption** | | | | | | | | | | | | | |
| **MCS** | 45.1 | 47.3 | 47.1 | 43.5 | 47.4 | 45.9 | 46.6 | -- | -- | -- | -- | -- | -- |
| **ALSPAC-G1** | 45.4 | 52.3 | 49.3 | 53.8 | 50.2 | 50.3 | -- | 50.2 | -- | -- | -- | -- | -- |
| **NS** | 41.7 | 49.6 | 47.6 | 44.4 | 47.3 | 46.2 | -- | 46.8 | -- | -- | -- | -- | -- |
| **BCS 70** | 46.9 | 50.7 | -- | -- | 49.0 | 49.2 | -- | -- | -- | 49.1 | -- | -- | -- |
| **NCDS** | 42.8 | 35.7 | -- | -- | 40.4 | 37.4 | -- | -- | -- | -- | 39.0 | -- | -- |
| **NSHD** | 11.7 | 9.3 | -- | -- | 9.7 | 12.0 | -- | -- | -- | -- | -- | 10.4 | -- |
| **USOC** | 50.7 | 52.4 | 48.6 | 50.7 | 51.6 | 56.4 | 52.7 | 66.9 | 66.2 | 66.4 | 57.5 | 22.1 | 9.9 |
| **ELSA** | 33.9 | 26.9 | 29.4 | 41.5 | 29.1 | 34.4 | -- | -- | -- | 51.6 | 46.7 | 20.8 | 8.0 |
| **GS** | 21.8 | 20.3 | 20.7 | 36.4 | 20.3 | 21.4 | -- | 23.6 | 31.9 | 34.9 | 27.7 | 9.0 | 4.1 |
| **ALSPAC-G0** | 50.7 | 48.0 | 48.7 | 49.5 | 48.9 | 47.9 | -- | -- | -- | 50.2 | 50.0 | 38.0 | 11.8 |

| | | | | | | | | | | | | | |
|---|---|---|---|---|---|---|---|---|---|---|---|---|---|
| **TWINSUK** | 35.9 | 30.4 | 30.9 | 29.5 | 26.3 | 35.1 | 50 | 37.3 | 44 | 42.3 | 40.7 | 25.2 | 20 |
| **GLAD** | 34.6 | 43.4 | 41.5 | 51.3 | 42.8 | 41.2 | 65.2 | 46.9 | 41.2 | 39.4 | 32.9 | 11.7 | 12.2 |
| **Any housing disruption** | | | | | | | | | | | | | |
| **MCS** | 31.8 | 34.3 | 35.1 | 23.4 | 26.5 | 38.9 | 33.5 | -- | -- | -- | -- | -- | -- |
| **ALSPAC-G1** | 24.3 | 22.8 | 23.3 | 24.0 | 21.6 | 30.0 | -- | 23.3 | -- | -- | -- | -- | -- |
| **NS** | 13.5 | 14.4 | 14.2 | 13.9 | 13.1 | 15.1 | -- | 14.1 | -- | -- | -- | -- | -- |
| **BCS 70** | 11.9 | 15.0 | -- | -- | 13.9 | 13.4 | -- | -- | -- | 13.7 | -- | -- | -- |
| **NCDS** | 9.7 | 12.2 | -- | -- | 8.7 | 13.8 | -- | -- | -- | -- | 11.1 | -- | -- |
| **NSHD** | 2.4 | 3.8 | -- | -- | 3.5 | 2.3 | -- | -- | -- | -- | -- | 3.1 | -- |
| **USOC** | 29.6 | 33.9 | 31.9 | 31.0 | 31.1 | 33.1 | 38.1 | 38.9 | 27.8 | 33.9 | 33.0 | 26.1 | 21.1 |
| **ELSA** | 24.5 | 24.4 | 23.5 | 37.3 | 22.9 | 30.1 | -- | -- | -- | 38.9 | 32.4 | 18.2 | 13.4 |
| **GS** | 5.9 | 8.7 | 7.7 | 9.1 | 5.8 | 9.8 | -- | 8.5 | 5.9 | 12.6 | 10.5 | 4.2 | 4.8 |
| **ALSPAC-G0** | 10.6 | 16.8 | 15.4 | 4.7 | 14.6 | 18.3 | -- | -- | -- | 17.9 | 15.1 | 11.8 | 0 |
| **TWINSUK** | 4.8 | 7.1 | 6.8 | 9.1 | 4.2 | 9.1 | 25 | 9.3 | 11 | 11.9 | 9.4 | 4.5 | 3.9 |
| **GLAD** | 10.4 | 12.8 | 12.2 | 15.9 | 13.1 | 11.8 | 28.8 | 12.1 | 7.2 | 11.5 | 9.2 | 6.9 | 5.8 |

Sources: MCS (Millennium Cohort Study); ALSPAC G1 (Children of the Avon Longitudinal Study of Parents and Children); NS (Next Steps); BCS 70 (1970 British Cohort Study), NCDS (National Child Development Study); NSHD (National Survey of Health and Development); USOC (Understanding Society); ELSA (English Longitudinal Study of Ageing); GS (Generation Scotland: the Scottish Family Health Study); TWINS UK (UK Adult Twin Registry); GLAD (Genetic Links to Anxiety and Depression), ALSPAC G0 (parents of ALSPAC). Weighted data. Notes: -- (not available/applicable).

#### Supplementary Table S6. Meta-regression assessing moderation by time since pre-pandemic mental health measure

| | Any Healthcare disruption | | Any economic disruption | | Any housing disruption | |
|---|---|---|---|---|---|---|
| | OR (95% CI) | p-value | OR (95% CI) | p-value | OR (95% CI) | p-value |
| **Time since pre-pandemic measure** | 0.99 (0.93, 1.05) | 0.63 | 1.03 (0.98, 1.08) | 0.26 | 0.99 (0.96, 1.03) | 0.84 |

**Table S7. Meta-analysed associations between standardised psychological distress and overall healthcare, economic and housing disruptions stratified by sex, education, ethnicity and age.**

| | **Healthcare disruption** | | **Economic disruption** | | **Housing disruption** | | **1 disruption vs none** | | **2+ disruptions vs none** | |
|---|---|---|---|---|---|---|---|---|---|---|
| | OR (95% CI) | $I^2$ | OR (95% CI) | $I^2$ | OR (95% CI) | $I^2$ | OR (95% CI) | $I^2$ | OR (95% CI) | $I^2$ |
| Female | 1.39 (1.28, 1.50) | 72.4% | 1.04 (0.96, 1.13) | 84.8% | 0.99 (0.96, 1.03) | 0.0% | 1.08 (1.02, 1.15) | 70.7% | 1.22 (1.13, 1.32) | 67.7% |
| Male | 1.44 (1.30, 1.60) | 52.4% | 1.03 (0.94, 1.14) | 69.4% | 1.01 (0.95, 1.07) | 0.0% | 1.08 (1.02, 1.14) | 17.3% | 1.29 (1.18, 1.40) | 34.5% |
| Degree | 1.41 (1.26,1.58) | 72.0% | 1.10 (1.03, 1.19) | 66.9% | 1.01 (0.97, 1.06) | 0.0% | 1.06 (0.99, 1.13) | 58.8% | 0.84 (0.56, 1.25) | 97.9% |
| No degree | 1.40 (1.29, 1.51) | 63.5% | 1.01 (0.92, 1.10) | 83.3% | 0.99 (0.95, 1.03) | 0.0% | 1.08 (0.99, 1.17) | 82.0% | 1.20 (1.10, 1.31) | 64.8% |
| White | 1.42 (1.28, 1.57) | 82.4% | 1.04 (0.93, 1.16) | 90.5% | 1.00 (0.96, 1.03) | 0.0% | 1.08 (1.00, 1.17) | 80.8% | 1.23 (1.12, 1.36) | 79.7% |
| Ethnic minority | 1.53 (1.17, 2.00) | 54.4% | 1.04 (0.85, 1.27) | 59.1% | 1.13 (0.97, 1.31) | 22.0% | 1.10 (0.93, 1.30) | 32.0% | 1.36 (1.12, 1.65) | 27.9% |
| 16-24 years | 1.42 (1.24, 1.62) | 0.0% | 1.05 (0.86, 1.29) | 82.5% | 0.99 (0.85, 1.17) | 70.3% | 1.00 (0.91, 1.10) | 0.0% | 1.13 (0.89, 1.44) | 78.0% |
| 25-34 years | 1.50 (1.16, 1.95) | 52.4% | 1.11 (1.01, 1.23) | 28.7% | 1.07 (0.97, 1.18) | 0.0% | 1.09 (0.94, 1.26) | 54.6% | 1.29 (1.15, 1.44) | 0.0% |
| 35-44 years | 1.70 (1.46, 1.98) | 0.0% | 0.97 (0.73, 1.30) | 88.1% | 1.17 (0.81, 1.69) | 82.3% | 0.99 (0.89, 1.10) | 21.2% | 1.27 (0.96, 1.67) | 65.2% |
| 45-54 years | 1.53 (1.39, 1.68) | 22.0% | 0.95 (0.86, 1.04) | 58.3% | 0.93 (0.87, 0.99) | 0.0% | 1.05 (0.94, 1.19) | 66.7% | 1.18 (0.97, 1.42) | 77.3% |
| 55-64 years | 1.42 (1.22, 1.65) | 81.5% | 0.96 (0.82, 1.11) | 90.3% | 0.98 (0.93, 1.04) | 0.0% | 1.07 (1.00, 1.15) | 50.5% | 1.17 (1.06, 1.29) | 48.1% |
| 65-74 years | 1.51 (1.13, 2.03) | 91.8% | 1.06 (0.90, 1.25) | 76.1% | 0.99 (0.91, 1.08) | 0.0% | 1.21 (1.12, 1.31) | 18.5% | 1.20 (1.03, 1.41) | 42.2% |
| 75+ years | 1.51 (1.17, 1.96) | 44.2% | 1.01 (0.78, 1.29) | 51.4% | 1.02 (0.86, 1.20) | 10.2% | 1.31 (1.00, 1.70) | 71.0% | 1.48 (1.01, 2.16) | 51.8% |

Adjusted for age, sex, ethnicity, education, and UK Nation
